# The Dynamics of Cognitive Decline towards Alzheimer’s Disease Progression: Results from ADSP-PHC’s Harmonized Cognitive Composites

**DOI:** 10.1101/2025.01.01.25319850

**Authors:** Kaidi Kang, Panpan Zhang, Logan Dumitrescu, Shubhabrata Mukherjee, Michael L. Lee, Seo-Eun Choi, Emily H. Trittschuh, Jesse Mez, Andrew J. Saykin, Katherine A. Gifford, Rachel F. Buckley, Xiaoting Gao, Jianing Di, Paul K. Crane, Timothy J. Hohman, Dandan Liu

**Affiliations:** Department of Biostatistics, Vanderbilt University Medical Center, 2525 West End Avenue, Suite 1100, Nashville, TN 37203, USA; Vanderbilt Memory & Alzheimer’s Center, Vanderbilt University Medical Center, 3319 West End Avenue, 8th Floor, Nashville, TN 37203, USA; Vanderbilt Genetics Institute, Vanderbilt University Medical Center, 1211 Medical Center Dr, Nashville, TN 37232, USA; Department of Neurology, Vanderbilt University Medical Center, 1211 Medical Center Dr, Nashville, TN 37232, USA; Department of Medicine, University of Washington, 1959 NE Pacific St, Seattle, WA 98195, USA; Department of Psychiatry and Behavioral Sciences, University of Washington School of Medicine, 1959 NE Pacific Street, Seattle, WA 98195-6560, USA; Geriatric Research Education Clinical Center, VA Puget Sound Health Care System, 1660 S Columbian Way, Seattle, WA 98108, USA; Department of Neurology, Boston University School of Medicine, 85 East Concord Street, 1st floor Boston, MA 02118, USA; Department of Radiology and Imaging Sciences, Indiana University School of Medicine, 550 N. University Blvd, Room 0663, Indianapolis, IN 46202, USA; Department of Anatomy & Neurobiology, Boston University School of Medicine, 72 East Concord St, Boston, MA 02118, USA; Department of Neurology, Massachusetts General Hospital and Harvard Medical School, 55 Fruit Street Boston, MA 02114, USA; Center for Alzheimer Research and Treatment, Department of Neurology, Brigham and Women’s Hospital, 75 Francis Street, Boston MA 02115, USA; Melbourne School of Psychological Science, University of Melbourne, Grattan Street, Parkville, VIC 3010, Australia; Janssen China Research & Development, 65 Gui-Qing Rd, Shanghai 200233, China

**Keywords:** ADSP-PHC, harmonized composite measures, sigmoidal mixed model, temporal order, cognitive decline

## Abstract

**INTRODUCTION:** Accurately assessing temporal order of cognitive decline across multiple domains is critical in Alzheimer’s disease (AD). Existing literature presented controversial conclusions likely due to the use of a single cohort and different analytical strategies.

**METHODS:** Harmonized composite cognitive measures in memory, language and executive functions from 13 cohorts in the ADSP-PHC data are used. A novel double anchoring events-based sigmoidal mixed model was developed using time to the incident of AD diagnosis as the time scale.

**RESULTS:** Decline in memory occurred before decline in language which was followed by the decline in executive function. Throughout the entire AD continuum, APOE-*ε*4 non-carriers and non-Hispanic Whites showed better memory performance, respectively, in all three cognitive domains.

**DISCUSSION:** Using harmonized data across multiple cohorts is the key to accurately characterizing the temporal order of AD biomarkers. Time to incident AD diagnosis should be used as the time scale for reproducibility purposes.

## 1 Background

Accurately assessing the temporal order of cognitive decline across multiple domains is critical for neuropsychologists to develop cognitive tests and batteries that are more sensitive to changes in pre-clinical AD^1^. However, existing literature presents controversial conclusions. Some studies suggest that early-stage AD affects memory first with a relatively minor impact on speech skills^2^, whereas some other studies find that language performance deficits can appear early before impairment in episodic memory^3^. In addition, there are studies concluding that impaired memory and executive function are the earliest-observable hallmarks of AD^4^ and that executive dysfunction occurs earlier than language disturbance and the decline in other cognitive skills ^5^. On the contrary, there are also studies showing that memory loss and language impairment may concurrently arise at an early stage of AD, while executive functioning decline usually takes place last ^6,7^. These controversial conclusions are likely due to the use of a single cohort with underlying heterogeneity across studies and different analytical strategies.

In response to the emerging needs of the AD research community to have a perpetually curated long-lived legacy dataset, the Alzheimer’s Disease Sequencing Project Phenotype Harmonization Consortium (ADSP-PHC) was established in 2021 to harmonize the rich endophenotype data across cohort studies involved in Alzheimer’s Disease Sequencing Project (ADSP). To date, ADSP-PHC has harmonized cognitive domains for more than 80,000 participants and 323,267 visits from 20 cohorts, which provides valuable data resources to characterize and compare the temporal order of cognitive decline.

Various analytical strategies have been used to investigate cognitive decline relative to AD progression. When the focus of interest lies in the comparison of their temporal order, analytical strategies that align with the remarkable pathological cascade model are often considered^8,9^. This hypothetical model assumes AD-related markers follow a sigmoidal shape (i.e., an “S” shape) as AD clinical stages progress and has engendered a large body of studies that exploit sigmoidal mixed models (SMM) to investigate the dynamics of AD-related markers ^10–15^. Nonetheless, there are still some challenges in the application of SMM for characterizing the dynamics of AD-related markers. First, AD progression is a slow process with many AD-related markers becoming abnormal at a preclinical stage when participants still have normal cognition (NC) or experience mild cognitive impairment (MCI)^16–20^, followed by AD symptom onset decades afterward. An ideal dataset should consist of longitudinal measurements from a large representative cohort with decades of follow-up beginning in middle age. However, collecting such a dataset is extremely costly and almost unfeasible. Second, existing literature assessing the dynamics of AD-related markers lacks a consensus time scale (i.e., the horizontal axis), making it extremely difficult to reproduce and consolidate scientific results across multiple studies. Some studies used latent trait descriptors of the underlying AD pathophysiological process as the time scale^21,22^. However, this approach has very low reproducibility as the quantification of AD progression scale is often study-specific, thus limiting its general applicability. Other studies chose to use the time to an anchoring event such as incident clinical diagnosis of AD^23,24^. Although such a time scale is clinically meaningful, this approach excludes participants without an observed incident clinical diagnosis of AD (i.e., the anchoring event) and thus results in information loss and potential selection bias.

Our work aims to characterize the cognitive decline in three domains (memory, language, and executive function) and compare their temporal order over the entire AD continuum, using harmonized data from multiple cohorts from ADSP-PHC. A novel double anchoring events-based sigmoidal mixed model (DSMM) was developed to address the aforementioned challenges related to reproducibility, selection bias, and cohort heterogeneity. Such methodology advancement allows the inclusion of participants without observed incident AD diagnosis but with observed incident MCI diagnosis into modeling the dynamics of cognitive decline toward AD onset, and thus naturally includes and connects different segments of AD progression continuum from participants to form a unified continuum for an aging population.

## 2 Methods

### 2.1 Harmonized ADSP-PHC Cognitive Data

Each fall, ADSP-PHC collects data from cohorts involved in ADSP and conducts data curation and harmonization. Since neuropsychological protocols vary across studies, ADSP-PHC applied bifactor models to harmonize and co-calibrate cognitive composite scores for memory, language, and executive function across multiple AD cohorts^25^. Higher values of the cognitive composite scores indicate better cognitive performance. The current study used the 2023 freeze data and included harmonized cognitive scores from 13 longitudinal cohorts. Namely, they are the Adult Change in Thought (ACT)^26^, the Alzheimer’s Disease Neuroimaging Initiative (ADNI)^27^, the Biomarkers for Older Controls at Risk for Dementia (BIOCARD) cohort^28^, the Baltimore Longitudinal Study of Aging (BLSA)^29^, the Estudio Familiar de Influencia Genetica en Alzheimer (EFIGA) cohort, the National Alzheimer’s Coordinating Center (NACC)^30^, the Rush Memory and Aging Project (MAP)^31,32^, the Minority Aging Research Study (MARS)^33^, the National Institute on Aging Late Onset of Alzheimer’s Disease (NIA-LOAD) cohort, the Religious Orders Study (ROS)^32^, the Memory and Aging Project from the Knight Alzheimer’s Disease Research Center at the Washington University in St. Louis (WASHU), the Wisconsin Registry for Alzheimer’s Prevention (WRAP)^34^, and the Washington Heights/Inwood Columbia Aging Project (WHICAP)^35,36^.

Modest exclusion criteria were applied to maximize the use of all available data (Figure 1).

**Figure 1:**
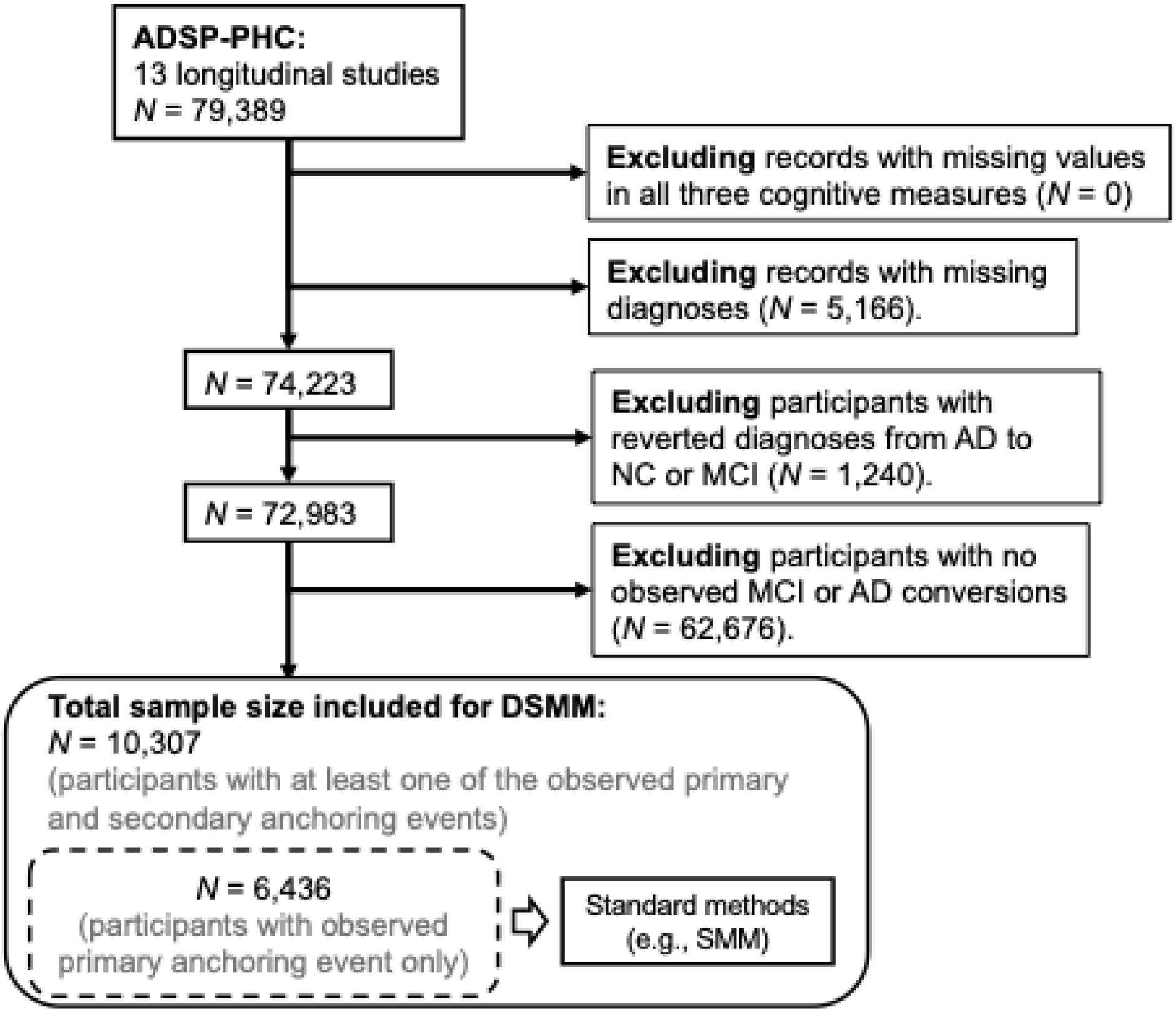
Participant Inclusion and Exclusion Flowchart for the DSMM Analysis. After removing records missing all three cognitive scores, records with missing diagnoses, participants with reverted diagnoses, and participants without observed MCI or AD conversion were excluded. A total of 10,307 participants were included in the DSMM analysis. Among them, only 6,436 participants have observed incident AD diagnosis and could be used in the conventional SMM analysis.

Specifically, observations missing all three cognitive scores (i.e., memory, language, and executive function) were excluded, followed by the exclusion of the observations with missing clinical diagnoses. Then, participants with reverted diagnoses after an AD diagnosis (i.e., receiving NC and/or MCI after AD diagnosis) were excluded. Lastly, participants with only one diagnosis throughout their follow-ups (e.g., NC, MCI, or AD) were excluded.

### 2.2 Double anchoring events-based Sigmoidal Mixed Model

This study uses time to incident clinical diagnosis of AD (*t_AD_*) as the time scale, where the primary anchoring event of incident AD diagnosis is marked as time 0. All longitudinal measurements are properly aligned on this new time scale, and thus visits before the incident clinical diagnosis of AD have negative values of *t_AD_* . For subject *i* at time *t_AD_*, a harmonized cognitive outcome *Y_i_*>(*t_AD_*) under the three-parameter SMM^37^ can be specified as

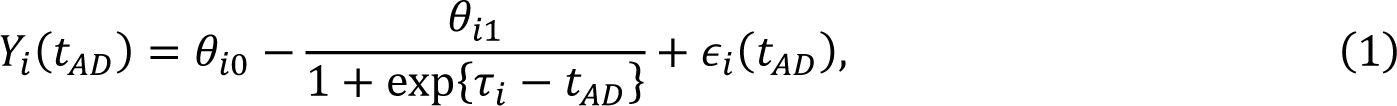

where θ_*i*0_, θ_*i*1_ and τ_*i*_ denote subject-specific trajectory parameters that follow a multivariate normal distribution with mean θ_0_, θ_1_ and τ, respectively. Specifically, θ_0_ is the population averaged cognitive measure prior to cognitive decline; θ_1_ is the population averaged cognitive decline over the entire AD progression with positive values with larger values denoting greater decline; τ is the population averaged time for the cognitive outcome to decline by half (i.e., half-life) with negative values denoting half decline prior to incident AD diagnosis. By convention, ϵ_*i*_ (*t_AD_*) is the error term assumed to follow a normal distribution. The SMM (1) allows subject-specific parameters to precisely capture the heterogeneity of individual trajectories.

Although the aforementioned data challenge to investigate temporal order of cognitive decline could be remarkably addressed by leveraging ADSP-PHC’s harmonized cognitive measures from multiple cohorts, the existing SMM approach still requires observed *t_AD_*, which means only participants with observed incidence of AD diagnosis could be included in the analysis, resulting in potential selection bias. To overcome this limitation, we proposed a novel approach to integrate the incidence of MCI diagnosis as a secondary anchoring event in the formation of *t_AD_*. Let *t_MCI_* denote time to incidence MCI diagnosis and let *D*_*i*_ = *t_MCI_* − *t_AD_* denote the duration between incident MCI and AD diagnoses. For participants with observed *t_MCI_*, but not observed *t_AD_*, the SMM model can be revised as

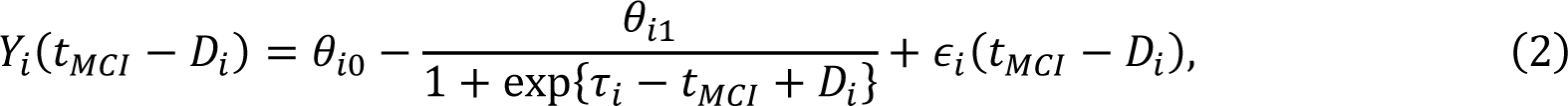

By including *D*_*i*_, the time scale *t_MCI_* can be realigned to *t_AD_*, allowing the inclusion of participants with either of the anchoring events observed into the characterization of cognitive decline and temporal order comparison. Although *D*_*i*_ is also not observed, it could be parameterized in a similar fashion as the trajectory parameters and simultaneously estimated in the overall modeling process. Combining models (1) and (2), the proposed DSMM is formulated as

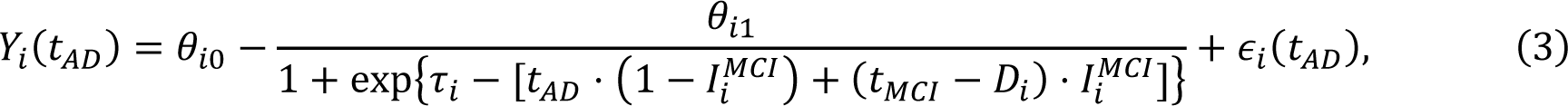

where *I^MCI^_i_* is an indicator variable with a value of 1 for participants with observed incidence MCI diagnosis and 0 otherwise.

Figure 2 demonstrates the conceptual framework of the extension from SMM to the proposed DSMM using the ADSP-PHC’s longitudinal harmonized memory composite measure. In the first step, longitudinal outcomes from participants with observed incident AD diagnosis are aligned on time to incident AD diagnosis *t_AD_* (Figure 2a). Next, longitudinal outcomes from participants with observed incident MCI diagnosis are aligned on the time to incident MCI diagnosis *t*_MCI_ (Figure 2b). Lastly, observations in Figure 2b are shifted by the subject-specific duration between the incident MCI and AD diagnosis *D*_*i*_ such that all longitudinal measures are aligned on *t_AD_* (Figure 2c). Participants with both incident AD diagnosis and incident MCI diagnosis observed can be included in either Figure 2a or Figure 2b. Investigation on the likelihood function suggests including these participants in Figure 2b will increase information used to estimate *D*_*i*_, and thus will improve the overall precision of parameter estimates. Therefore, we recommend including these participants in Figure 2b which is consistent with the use of *I^MCI^_i_* in model (3).

**Figure 2:**
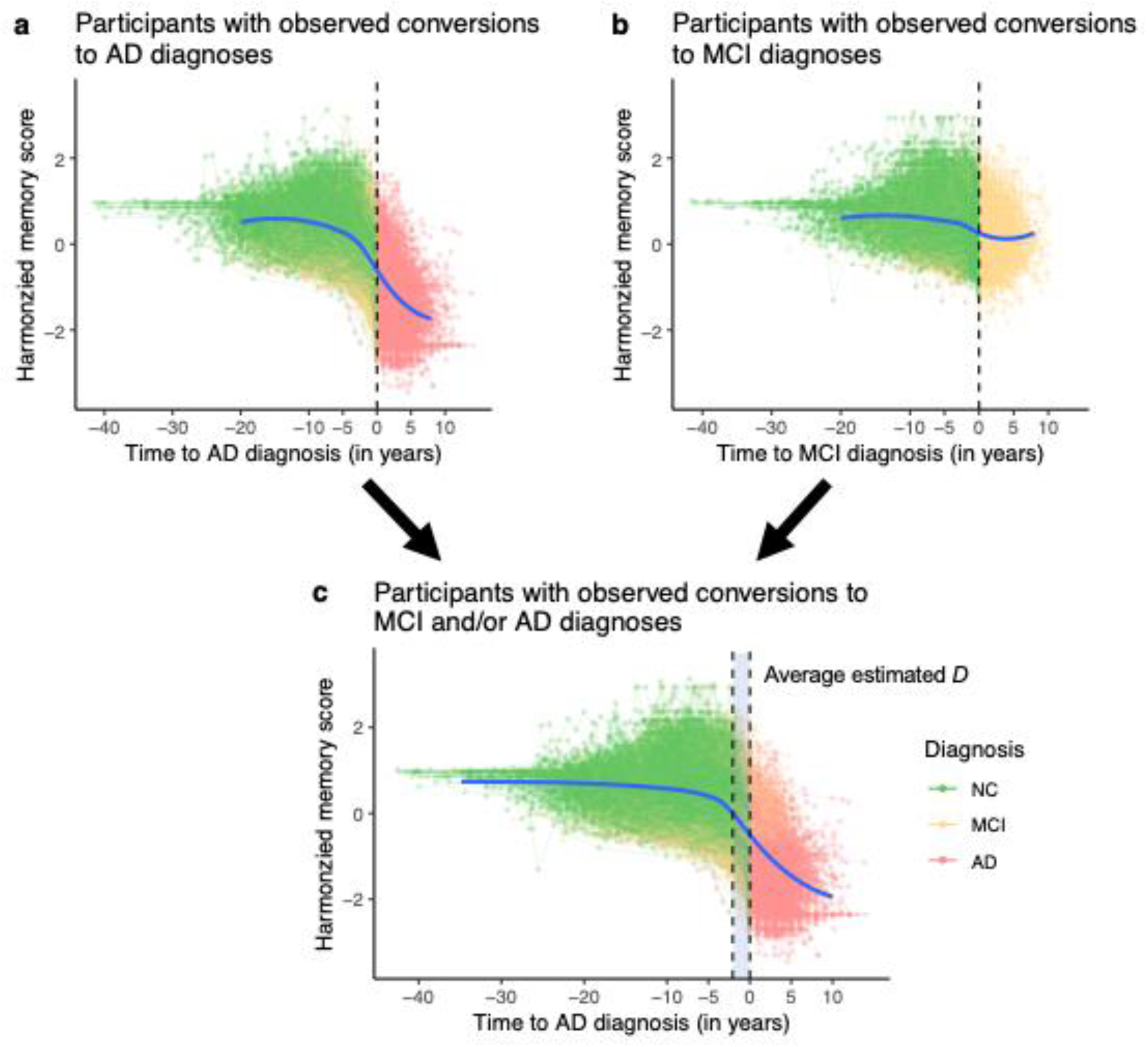
Conceptual Model Framework of the Proposed DSMM. Alignment of the primary and secondary anchoring event time scales using the ADSP-PHC’s harmonized memory composite measures. (a) Longitudinal measures on the time scale of “time to the primary anchoring event” for participants with observed primary anchoring event (i.e., incident AD diagnosis); (b) Longitudinal measures on the time scale of “time to the secondary anchoring event” for participants with observed secondary anchoring event (i.e., incident MCI diagnosis); (c) The secondary anchoring event time scale is shifted by a subject-specific measure of duration between the two (potential) anchoring events *D_i_* such that longitudinal measures from participants with observed secondary anchoring events could be included to characterize the dynamics of cognitive decline over the time scale of “time to the primary anchoring event.” The blue curves are the locally estimated scatterplot smoothing (LOESS) curves.

It is worth noting that the proposed DSMM is flexible in the sense that it could not only be used as an unadjusted model that provides estimates for each of the parameters (θ_0_, θ_1_ and τ and *D*) but also be used as an adjusted model allowing each parameter dependent on some covariates. In addition, the proposed DSMM could also account for heterogeneity across multiple cohorts by introducing nested random effects since it falls in a broad class of nonlinear mixed effects models^38^. Any existing statistical programming software (e.g., R) that could fit nonlinear mixed effect models could be used for analysis.

### 2.3 Statistical Analyses

The proposed DSMM was first applied to characterize the overall trajectory of decline in each cognitive domain, where the parameters were not adjusted for any covariates. A hierarchical structure of random effects with participants nested within cohorts was used to account for heterogeneity across multiple cohorts in ADSP-PHC. Temporal orders of decline were assessed by comparing the half-life *τ* for each cognitive domain. Under the three-parameter SMM, a smaller *τ* indicates an earlier decline. Moreover, this comparison was repeated within each single cohort to explore the influence of cohort heterogeneity on the conclusion of temporal orders. Within these single-cohort analyses, only subject-level random effects were included. Furthermore, adjusted analyses were conducted to compare trajectories across different genetic risk, gender, and race/ethnicity groups. Specifically, DSMM was applied by allowing all trajectory parameters to be linearly dependent on APOE-*ε*4 carrier status, gender, and race/ethnicity.

For all analyses, logarithm transformation was imposed on *Di* to accommodate its natural constraints, as it should always be positive. To allow sufficient precision in estimating *Di* and thus reliable time shift from *t*_MCI_ to *t_AD_*, a linear model for log(*Di*) was integrated into DSMM covarying for age at the happening of the secondary anchoring event (centered at the empirical average of 77 years old), gender, APOE-*ε*4 carrier status, and two-way interactions between APOE-*ε*4 carrier status and all other factors. Cubic splines with two degrees of freedom were added to the centered age allowing for non-linearity.

In practice, convergence issues often arise when fitting DSMM using a frequentist approach, due to non-linearity, large sample size, and data heterogeneity. To circumvent this challenge, we resorted to a Bayesian algorithm by utilizing the built-in functions from the well-developed brms package^39^. Informative priors were generated based on the estimates of the reduced SMMs. Convergence was verified using standard diagnostic plots. Credible intervals (CIs) and posterior predictive *p*-values were reported for inference.

These two Bayesian-specific statistics are conceptually similar to confidence intervals and *p*-values in frequentist inference. All statistical analyses were conducted in R (Version 4.3.2). The example codes for model fitting are available on GitHub at https://github.com/KaidiK/DSMM.

## 3 Results

### 3.1 Data Summary

The 2023 freeze ADSP-PHC data were used, which consists of 13 longitudinal AD cohorts. After applying the pre-specified exclusion criteria, a total of 63,309 longitudinal records from 10,307 participants were included in the analysis. Among them, 3,871 (37.6%) participants would have been excluded if the conventional SMM was used instead of the newly developed DSMM, as they only have observed incident MCI diagnosis but not observed incident AD diagnosis.

Included participants from the 13 cohorts had a mean baseline age of 76.0 years, an average follow-up of 7.61 years, 60.1% female, 70.5% non-Hispanic Whites, and 39.4% APOE-*ε*4 carriers. See Table 1 for a summary of participant characteristics for the analysis data.

**Table 1:** Summary of participant characteristics of the ADSP-PHC data in the present analysis.

| Participant characteristics | *N* = 10,307 |
| --- | --- |
| **Gender** |  |
| Female | 6,190 (60.1%) |
| Male | 4,117 (39.9%) |
| **Baseline age** (in years) |  |
| Mean (SD) | 76.0 (8.01) |
| Median [Min, Max] | 76.0 [50, 110] |
| **Race/Ethnicity** |  |
| Non-Hispanic White | 7,262 (70.5%) |
| Others | 3,045 (29.5%) |
| **APOE-ε4 positive** |  |
| Yes | 3,661 (39.4%) |
| No | 5,639 (60.6%) |
| **Follow-up length** (in years) |  |
| Mean (SD) | 7.61 (5.59) |
| Median [Min, Max] | 6.24 [0.41, 42.90] |
| **Baseline harmonized memory score** |  |
| Mean (SD) | 0.23 (0.58) |
| Median [Min, Max] | 0.23 [−1.96, 2.94] |
| **Baseline harmonized language score** |  |
| Mean (SD) | 0.24 (0.60) |
| Median [Min, Max] | 0.25 [−1.90, 2.59] |
| **Baseline harmonized executive function score** |  |
| Mean (SD) | 0.04 (0.66) |
| Median [Min, Max] | 0.08 [−2.42, 2.42] |
| **Baseline diagnosis** |  |
| NC | 6,598 (64.0%) |
| MCI | 3,709 (36.0%) |
| **Observed incident AD diagnosis** |  |
| Yes | 6,436 (62.4%) |
| No | 3,871 (37.6%) |

SD: Standard deviation

### 3.2 Overall Cognitive Trajectories

The overall trajectory parameter estimates from the unadjusted analysis using 13 cohorts data from ADSP-PHC were shown in Table 2. The population averaged initial scores (i.e., θ_0_) were different across the three cognitive domains, where the highest was memory 0.34 (95% CI: [0.16, 0.50]) and the lowest was executive function 0.20 (95% CI: [*−*0.05, 0.41]). In addition, the memory score had the greatest population averaged decline of 1.42 (95% CI: [1.23, 1.63]), followed by language 1.20 (95% CI: [1.01, 1.40]), whereas executive function had the least decline 1.15 (95% CI: [0.98, 1.31]). Half declines in the memory domain took an average of 0.78 years (95% CI: [*−*1.16*, −*0.38]) before incident clinical diagnosis of AD. Half decline in the language domain occurred around incident clinical diagnosis of AD (τ = *−*0.09 with 95% CI: [*−*0.45, 0.31]), whereas half declines in the executive function domain happened after the incident clinical diagnosis of AD (i.e., τ = 0.25 with 95% CI: [*−*0.04, 0.57]).

**Table 2:** Overall trajectory parameter estimates in the DSMM applied to the entire ADSP-PHC analysis data.

| | Estimate | SE | 95% CI | $p$ |
| --- | --- | --- | --- | --- |
| **Memory** |  |  |  |  |
| $\theta_0$ | 0.34 | 0.09 | (0.16, 0.50) | $< 0.01$ |
| $\theta_1$ | 1.42 | 0.10 | (1.23, 1.63) | $< 0.01$ |
| $\tau$ | -0.78 | 0.20 | (-1.16, -0.38) | $< 0.01$ |
| **Language** |  |  |  |  |
| $\theta_0$ | 0.24 | 0.09 | (0.07, 0.40) | $< 0.01$ |
| $\theta_1$ | 1.20 | 0.10 | (1.01, 1.40) | $< 0.01$ |
| $\tau$ | -0.09 | 0.20 | (-0.45, 0.31) | 0.60 |
| **Executive function** |  |  |  |  |
| $\theta_0$ | 0.20 | 0.12 | (-0.05, 0.41) | 0.09 |
| $\theta_1$ | 1.15 | 0.09 | (0.98, 1.31) | $< 0.01$ |
| $\tau$ | 0.25 | 0.16 | (-0.04, 0.57) | 0.10 |

SE: Standard error; CI: Credible Interval; $p$ : Posterior Predictive $P$ -value.

The estimates of τ suggested a temporal ordering of declines in the three cognitive domains (i.e., memory, followed by language, and then executive function), which was also graphically reflected by their fitted trajectories shown in Figure 3. Memory decline took place notably earlier than declines in language and executive function domains. Further, declines in language occurred slightly earlier than declines in executive functioning. Hence, the temporal order of cognitive declines in the three domains of interest is consistent with the order of the estimated values of *τ* from DSMM.

**Figure 3:**
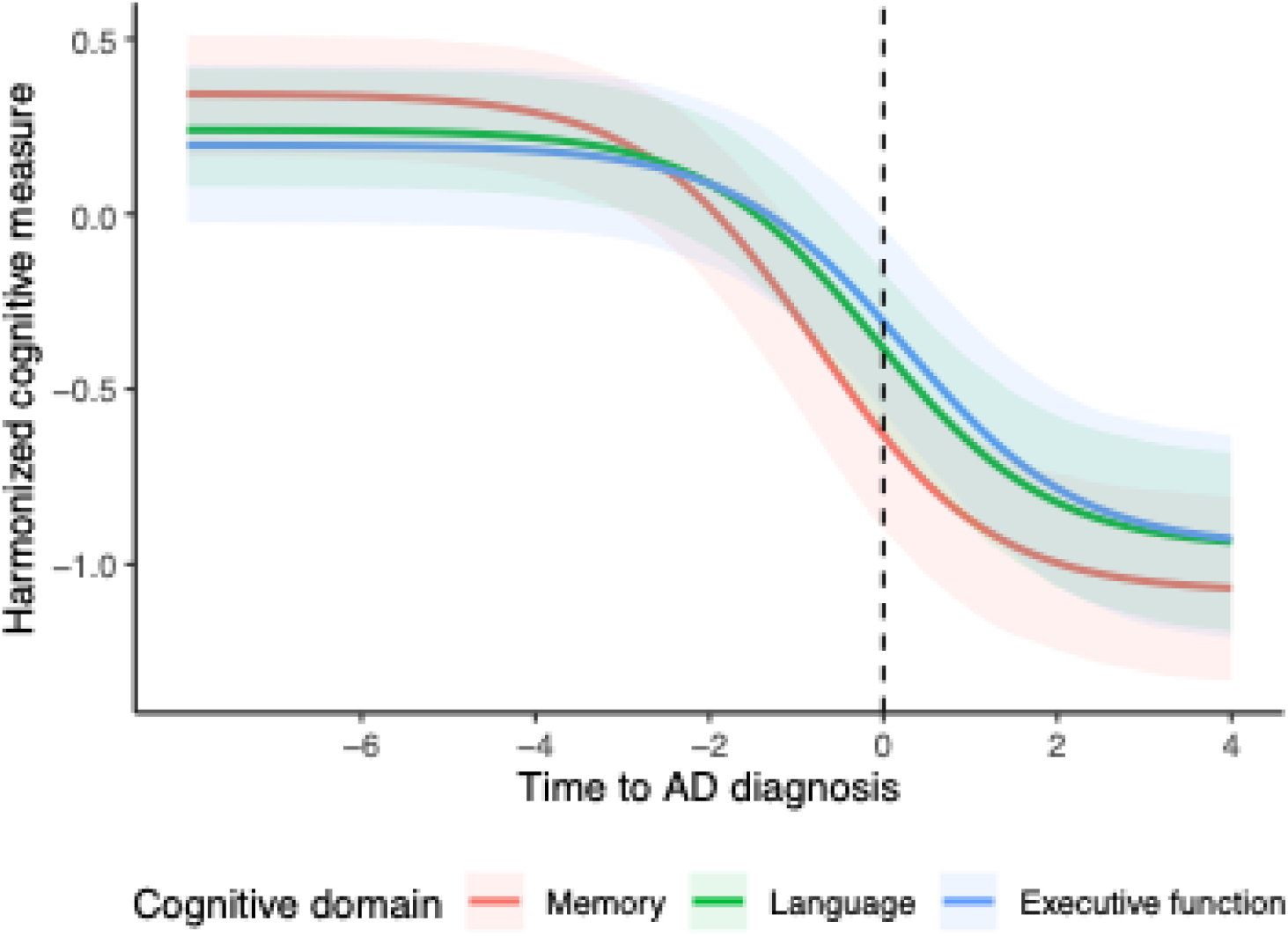
Overall trajectory from the unadjusted analyses. The fitted trajectories of three cognitive domains (i.e., memory, language, and executive function) from unadjusted DSMMs applied to 13 cohorts of ADSP-PHC data. The shaded areas are 95% confidence bands.

Results from single cohort DSMM analysis were quite different across studies. For each cognitive domain, there was remarkable heterogeneity in fitted trajectories across cohorts (Supplementary Figure S1). The temporal order of declines in the three cognitive domains also varied dramatically across single-cohort analyses (Supplementary Figure S2). For some cohorts (ACT, ADNI, MAP, NIA-LOAD, WASHU, and WRAP), the fitted trajectories clearly showed that memory declined earlier than language and executive function whereas for other cohorts, this pattern is not clear.

### 3.3 Covariate-Dependent Cognitive Trajectories

Covariate effects on trajectory parameters were presented in Table 3. APOE-*ε*4 carriers tended to have lower initial scores than non-carriers in memory (*p <* 0.01), but not in language (*p* = 0.24) or executive function (*p* = 0.50). In addition, APOE-*ε*4 carriers tended to experience greater declines than the non-carriers in all three cognitive domains (*p*-values *<* 0.01). The difference in the time taken for memory to decline by half between APOE-*ε*4 carriers and non-carriers was not significant (*p* = 0.72), but half declines in language and executive function occurred much later among APOE-*ε*4 carriers than non-carriers (*p*-values *<* 0.01).

**Table 3:** Covariate estimates on trajectory parameters in the DSMM applied to the entire ADSP-PHC analysis data.

|  | Memory |  |  |  | Language |  |  |  | Executive Function |  |  |  |
| --- | --- | --- | --- | --- | --- | --- | --- | --- | --- | --- | --- | --- |
|  | Est | SE | 95% CI | *p* | Est | SE | 95% CI | *p* | Est | SE | 95% CI | *p* |
| $\theta_0$ | | | | | | | | | | | | |
| Intercept | 0.44 | 0.08 | (0.27, 0.58) | < 0.01 | 0.35 | 0.07 | (0.20, 0.47) | < 0.01 | 0.28 | 0.09 | (0.09, 0.45) | 0.01 |
| APOE-ε4: Pos | -0.08 | 0.01 | (-0.10, -0.06) | < 0.01 | 0.01 | 0.01 | (-0.01, 0.03) | 0.24 | 0.01 | 0.01 | (-0.01, 0.03) | 0.50 |
| Gender: Male | -0.14 | 0.01 | (-0.16, -0.12) | < 0.01 | -0.06 | 0.01 | (-0.08, -0.04) | < 0.01 | -0.02 | 0.01 | (-0.04, 0.00) | 0.07 |
| Race: Others | -0.18 | 0.01 | (-0.21, -0.16) | < 0.01 | -0.34 | 0.01 | (-0.37, -0.31) | < 0.01 | -0.37 | 0.01 | (-0.40, -0.35) | < 0.01 |
| $\theta_1$ | | | | | | | | | | | | |
| Intercept | 1.44 | 0.10 | (1.26, 1.66) | < 0.01 | 1.22 | 0.10 | (1.04, 1.42) | < 0.01 | 1.14 | 0.08 | (0.99, 1.30) | < 0.01 |
| APOE-ε4: Pos | 0.11 | 0.02 | (0.07, 0.15) | < 0.01 | 0.23 | 0.02 | (0.19, 0.27) | < 0.01 | 0.25 | 0.02 | (0.20, 0.29) | < 0.01 |
| Gender: Male | -0.12 | 0.02 | (-0.16, -0.07) | < 0.01 | -0.06 | 0.02 | (-0.11, -0.02) | 0.02 | -0.02 | 0.02 | (-0.06, 0.03) | 0.42 |
| Race: Others | -0.17 | 0.03 | (-0.22, -0.11) | < 0.01 | -0.23 | 0.03 | (-0.29, -0.18) | < 0.01 | -0.16 | 0.03 | (-0.22, -0.12) | < 0.01 |
| $\tau$ | | | | | | | | | | | | |
| Intercept | -0.87 | 0.22 | (-1.27, -0.43) | < 0.01 | -0.35 | 0.23 | (-0.75, 0.15) | 0.14 | 0.05 | 0.16 | (-0.24, 0.41) | 0.77 |
| APOE-ε4: Pos | -0.02 | 0.07 | (-0.16, 0.10) | 0.72 | 0.74 | 0.08 | (0.60, 0.89) | < 0.01 | 0.69 | 0.09 | (0.53, 0.86) | < 0.01 |
| Gender: Male | 0.29 | 0.06 | (0.16, 0.41) | < 0.01 | 0.16 | 0.08 | (0.00, 0.31) | 0.05 | 0.07 | 0.09 | (-0.12, 0.23) | 0.42 |
| Race: Others | 0.17 | 0.08 | (0.02, 0.34) | 0.04 | 0.22 | 0.10 | (0.03, 0.40) | 0.02 | -0.21 | 0.11 | (-0.41, 0.01) | 0.06 |

Est: Estimate; SE: Standard error; CI: Credible interval; *p*: Posterior Predictive *P*-value.

Male participants tended to have lower initial cognitive scores (*p*-values *<* 0.01) and less declines (*p*-values *≤* 0.02) in memory and language than female participants, while there were no significant differences between males and females in the initial cognitive scores or declines for executive function (*p*-values *≥* 0.07). Additionally, the half decline in memory and language occurred later for males than females (*p*-values *≤* 0.05), but there were no significant sex differences in time to half declines of executive function (*p* = 0.42).

Non-Hispanic White participants tended to have higher initial cognitive scores for all three domains (*p*-values *<* 0.01), and their cognitive measures declined more than participants of other racial and ethnic groups (*p*-values *<* 0.01). Non-Hispanic White participants underwent earlier declines in memory and language than participants of other racial and ethnic groups (*p*-values *≤* 0.04), but later decline in executive function although the difference was not significant (*p* = 0.06). To visualize the temporal orders of declines in the three cognitive domains across various participant groups, fitted trajectories by APOE-*ε*4 status, sex, and race/ethnicity groups were presented in Figure 4.

**Figure 4:**
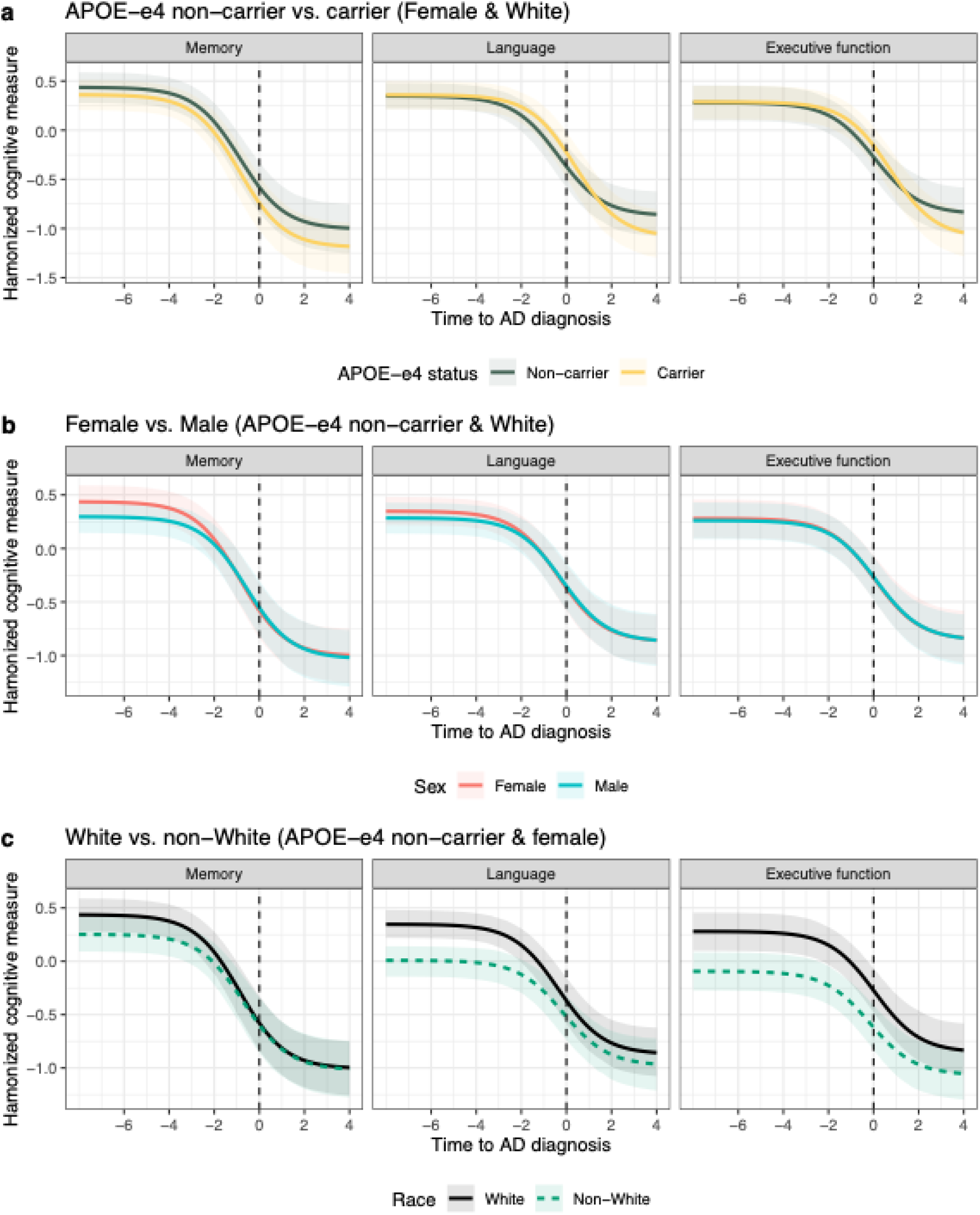
Covariate-adjusted fitted trajectories. The fitted trajectories of three cognitive domains (i.e., memory, language, and executive function) by (a) APOE-*ε*4 status: non-carrier vs. carrier, (b) Sex: Female vs. Male, and (c) Race/Ethnicity: Non-Hispanic White vs. Other. The shaded areas are 95% confidence bands.

## 4 Discussion

In this work, a novel DSMM was proposed to characterize longitudinal trajectories of cognitive markers over the continuum of AD progression with applications to harmonized cognitive measures from 13 cohorts in ADSP-PHC data. The proposed DSMM employed a time scale based on a secondary anchoring event in addition to the time scale based on generic anchoring events (e.g., incident clinical diagnosis of AD) to overcome the limitations of using time scales based on study-specific biomarkers^21^ and the challenges arising from varying participant entry points in standard longitudinal modeling. By considering secondary anchoring events (e.g., incident clinical diagnosis of MCI), DSMM provided a rigorous joint modeling framework for participants with either the primary or the secondary anchoring events to reduce inference bias caused by excluding participants with censored information about AD diagnosis in standard modeling procedures.

This work is among the first research exploring the temporal order of cognitive declines over the entire AD spectrum in various cognitive domains. ADSP-PHC plays a vital role by providing unique resources of harmonized cognitive measures from 13 longitudinal cohort studies. The results showed that cognitive changes in the memory domain exhibited the most extensive dynamic range than language and executive function. In addition, memory decline preceded the decline in the language domain, which preceded the decline in executive function. Interestingly, repeated analyses conducted in each cohort separately reveal different results. For example, results from BIO-CARD showed that executive function declined earlier than memory and language, but results from WASHU showed the cognitive decline in the language took place earliest. On the other hand, results from some other cohorts (ACT, ADNI, MAP, NIA-LOAD, WASHU, and WRAP) showed memory decline occurred earliest. Different results obtained from single cohort study partly explained why there were controversial conclusions in existing literature on the temporal orders of the three cognitive domains^2–7^. The controversial conclusions might be attributed to cohort heterogeneity (e.g., different recruitment targets and protocols), suggesting that a single cohort generally cannot represent the entire AD population, and further underscoring the value of unique harmonized data resources provided by ADSP-PHC.

Investigating trajectory parameter differences across demographic characteristics and genetic factors allowed us to compare the dynamics of cognitive decline across different groups. It should be noted that our analysis strategy using the time to incident AD diagnosis as the time scale under the SMM framework is quite different from conventional statistical approaches using the follow-up time as the time scale.

Therefore, our findings are not directly comparable to those from existing literature, especially when making inference on the rate of cognitive decline. Specifically, the estimates of the half-life trajectory parameter *τ* were used to compare the times to half cognitive decline. However, an earlier cognitive decline (smaller value of *τ*) does not necessarily mean faster cognitive decline because half-life depends on the initial cognitive performance, the magnitude of the decline and the rate of the decline. In conventional longitudinal analysis using follow-up time as the time scale, one can directly quantify and compare cognitive decline rates using the interactions between follow-up time and comparison groups of interest.

Our adjusted DSMM analyses suggested that APOE-*ε*4 carriers showed lower initial cognitive ability in memory than non-carriers, but such differences did not exist for language or executive function domains. In addition, APOE-*ε*4 carriers showed a greater decline in all three domains. Interestingly, half decline in language and executive function occurred later for APOE-*ε*4 carriers, but no difference was observed in time of half decline in memory between these two groups. Combining with the group-specific fitted trajectories, our findings suggested APOE-*ε*4 carriers had worse memory performance than non-carriers throughout the entire AD continuum. Although there was no difference in language and executive functions between APOE-*ε*4 carriers and non-carriers at the beginning of the decline, APOE-*ε*4 carriers tended to have worse cognitive performance in these two domains toward the later phase of the AD progression. Comparison between sexes suggested that female participants tended to have higher initial cognitive scores but experienced greater cognitive decline during AD progression and had an earlier half decline for memory and language than males. However, there was no difference between males and females in the dynamics of executive functions. These findings were generally consistent with existing literature that females generally do better in memory and language than males^40^, but have faster cognitive decline^41^. Comparison between race/ethnicity groups suggested that non-Hispanic Whites tended to have higher initial cognitive performance than non-Whites in all three cognitive domains. Although non-Hispanic Whites unexpectedly showed greater cognitive decline and an earlier half decline in all three cognitive domains than non-Whites, they still showed better cognitive performance throughout the entire AD continuum. This is consistent with existing literature^42^.

The current research has a few limitations which motivate our future studies. First, this research focuses on the characterization of longitudinal trajectories of cognitive markers toward AD progression such that the proposed DSMM was designed to only include participants presenting disease progression. However, it is common for participants to have one stable clinical diagnosis throughout the study period. The current version of DSMM is not yet able to handle these participants and thus an extension of the current DSMM is warranted. Second, competing risk of death potentially impacts the estimation precision and model reliability, therefore is necessary to consider in AD studies. Some basic concepts and methods discussed by Joseph *et al*.^43^ may be useful for future adaptions and extensions.

Despite these limitations, this study provides important new insights into the temporal order of declines in three cognitive domains by characterizing longitudinal trajectories toward AD progression. Future work will consider a larger and more representative sample as the participating cohorts of ADSP-PHC continue to grow. Besides, continued efforts should be made to consider more cognitive domains and other neurodegenerative markers related to AD progression for a comprehensive investigation. Continued exploration in this area is important to advance the understanding of the dynamics of AD biomarkers during AD progression.

## Data Availability

This secondary research work utilizes de-identified data obtained from primary data repositories. The data used in this study may be available upon reasonable request to the principal investigators of ADSP-PHC or the respective contributing studies.

## Abbreviations

AD: Alzheimer’s disease
MCI: Mild cognitive impairment
NC: Normal cognition
ADSP-PHC: The Alzheimer’s Disease Sequencing Project Phenotype Harmonization Consortium
APOE-*ε*4: Apolipoprotein E epsilon 4 allele
SMM: Sigmoidal mixed model
DSMM: Double anchoring events based sigmoidal mixed model
SD: Standard deviation
SE: Standard error

## Acknowledgments

ACT was supported by U19-AG0665677 and U01-AG006781. ADNI data collection was supported by U01-AG024904 (M. Weiner, PI). ADNI is funded by the National Institute on Aging, the National Institute of Biomedical Imaging and Bioengineering, and through generous contributions from the following: AbbVie, Alzheimer’s Association; Alzheimer’s Drug Discovery Foundation; Araclon Biotech; BioClinica, Inc.; Biogen; Bristol-Myers Squibb Company; CereSpir, Inc.; Cogstate; Eisai Inc.; Elan Pharmaceuticals, Inc.; Eli Lilly and Company; EuroImmun; F. Hoffmann-La Roche Ltd. and its affiliated company Genentech, Inc.; Fujirebio; GE Healthcare; IXICO Ltd.; Janssen Alzheimer Immunotherapy Research & Development, LLC.; Johnson & Johnson Pharmaceutical Research & Development LLC.; Lumosity; Lundbeck; Merck & Co., Inc.; Meso Scale Diagnostics, LLC.; NeuroRx Research; Neurotrack Technologies; Novartis Pharmaceuticals Corporation; Pfizer Inc.; Piramal Imaging; Servier; Takeda Pharmaceutical Company; and Transition Therapeutics. The Canadian Institutes of Health Research is providing funds to support ADNI clinical sites in Canada. Private sector contributions are facilitated by the Foundation for the National Institutes of Health (www.fnih.org). The grantee organization is the Northern California Institute for Research and Education, and the study is coordinated by the Alzheimer’s Therapeutic Research Institute at the University of Southern California. ADNI data was disseminated by the Laboratory for Neuro Imaging at the University of Southern California. BIOCARD data collection was supported by U19-AG033655. BLSA data collection was supported by the Intramural Research Program of the National Institute on Aging, National Institute of Health, Baltimore, MD. EFIGA data collection was supported by R01-AG067501 (Richard Mayeux, PI). NACC data collection was supported by U24-AG072122 (Walter A. Kukull, PI). MAP data collection and genotyping were supported by P30-AG72975, R01-AG15819, and R01-AG17917. MARS data collection was supported by RF1-AG22018 (Lisa L. Barnes, PI). NIA-LOAD data collection was supported by U24-AG026395. ROS data collection and genotyping were supported by P30-AG10161 (David A. Bennett, PI) and R01-AG15819 (David A. Bennett, PI). WASHU data collection was supported by P01-AG003991 (Leonard Berg, PI) and P30-AG066444 (John Morris, PI). WRAP data collection was supported by RF1-AG027161 (Sterling Johnson, PI). WHICAP data collection was supported by R01-AG16206.

## Conflicts

Dr. Dumitrescu reported grants from NIH during the study. Dr. Mez reported grants from NIH during the study and grants from the Department of Defense outside the submitted work. Dr. Saykin reported grants from NIH to Indiana University and support from multiple NIH grants during the conduct of the study as well as support from Avid Radiopharmaceuticals, a subsidiary of Eli Lilly (in-kind contribution of PET tracer precursor); Bayer Oncology (scientific advisory board); Eisai (scientific advisory board); Siemens Medical Solutions USA (dementia advisory board); National Heart, Lung, and Blood Institute (MESA observational study monitoring board); and Springer-Nature Publishing (editorial office support as editor-in-chief for Brain Imaging and Behavior) outside the submitted work. Dr. Hohman reported grants from NIH during the study and is a member of a scientific advisory board for Vivid Genomics outside the submitted work. Dr. Liu reported grants from NIH during the study. No other disclosures were reported.

## Funding sources

This research was supported by U24-AG074855 (Timothy Hohman, Michael Cuccaro, A. Toga, MPI). Dr. Hohman was also supported by R01-AG079142. Dr. Dumitrescu was supported by R01-AG073439. Dr. Mukherjee was supported by R01-AG082730. Dr. Mez was supported by R01-AG061028. Dr. Saykin was supported by P30-AG010133 and R01-AG019771. Dr. Gifford was supported by R01-AG062826. Dr. Buckley was supported by R01-AG079142. Dr. Crane was supported by U01-AG0006781, U19-AG066567, and R01-AG029672.

## Consent Statement

All contributing studies included in this work received ethical oversight from their respective institutions. This study constitutes secondary research, utilizing de-identified data obtained from primary data repositories. In accordance with NIH policy, this research does not qualify as human subject research, and therefore, obtaining consent from individual participants is not required.

## Authors’ contributions

KK designed and set up the research, performed the data analysis, and wrote the first draft of the manuscript under the supervision of PZ and DL, who were all responsible for this manuscript. SM, MLL, SC, EHT, JM, KAG, RFB, PKC, and TJH were involved in data acquisition and generation. TJL played a lead role in funding acquisition and a supporting role in resources and writing of review and editing. KK, PZ, DL, XG and JD provided critical input on the method development. EHT, JM, RFB, TJH, PZ, and DL provided input on the data analysis and interpretation. All authors read and approved the final manuscript.

## Supplementary Figures Legends

**Figure S1:**
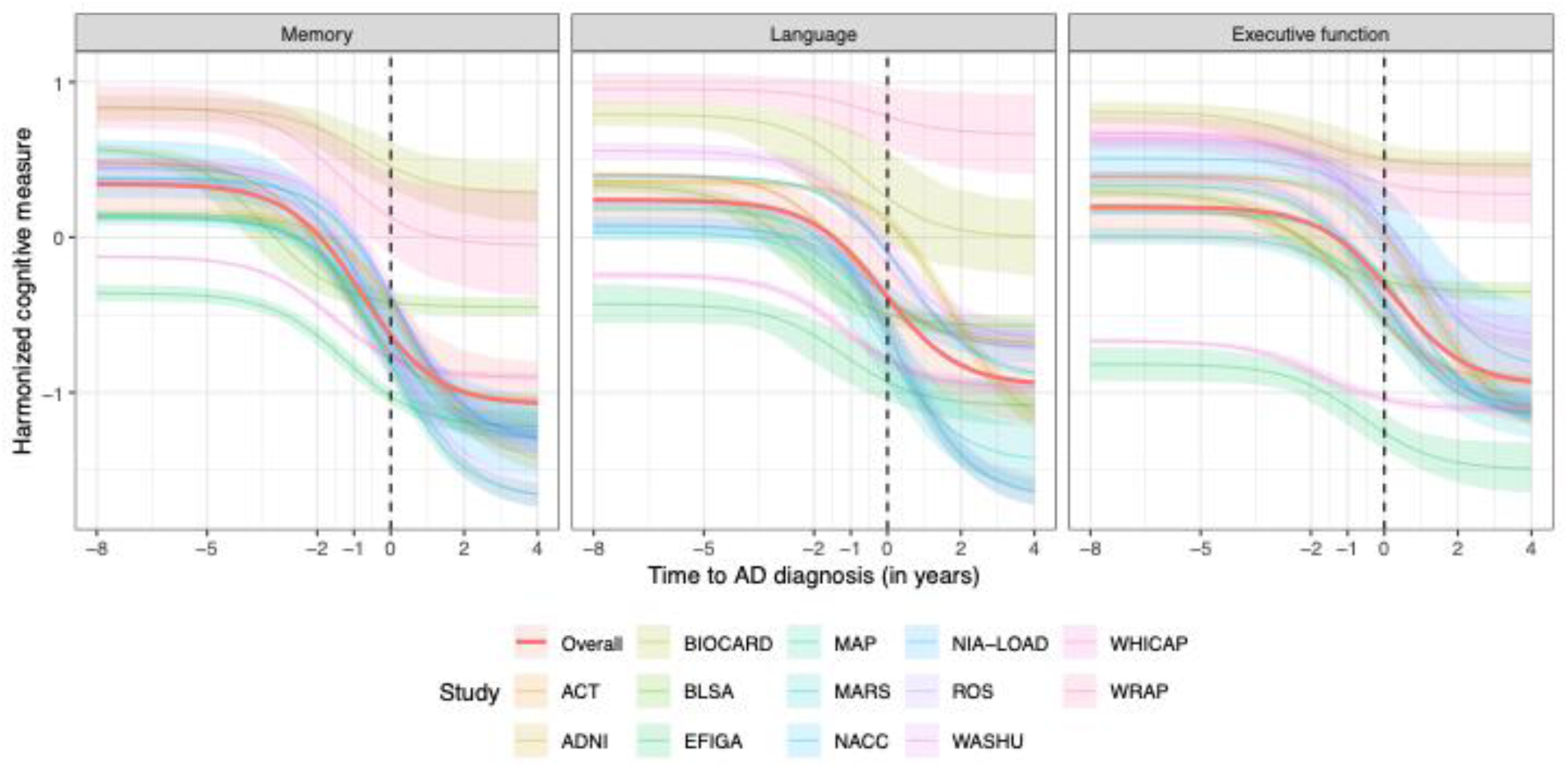
The estimated study-specific and overall trajectories of different cognitive domains (i.e., memory, language, and executive function) relative to the time to AD diagnosis. The overall trajectories (red) were fitted using the all data from the thirteen studies. Each study-specific trajectory for a cognitive domain was fitted using the corresponding study data. The shaded areas are 95% confidence intervals (CIs). For a cognitive domain, using data from different studies resulted in a quite amount of heterogeneity in the estimated trajectories.

**Figure S2:**
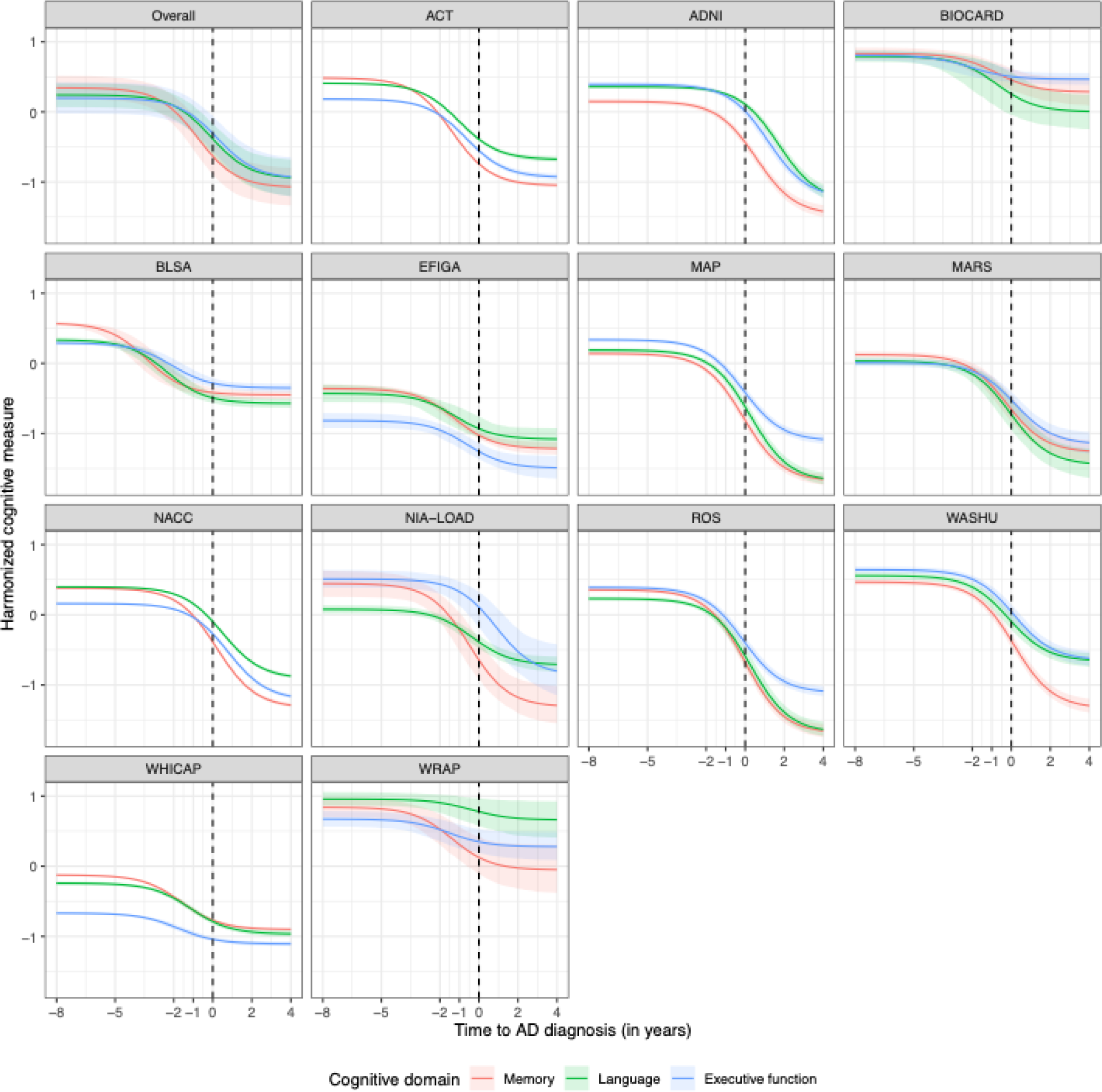
The estimated trajectories in the three cognitive domains (i.e., memory, language, and executive function) relative to the time to the diagnosis of AD in different studies. Each study-specific trajectory for a cognitive domain was fitted using the corresponding study data only. The shaded areas are 95% confidence intervals (CIs). Studying the temporal order of decline across cognitive domains only based on a single study may lead to different conclusions.

**Figure S3:**
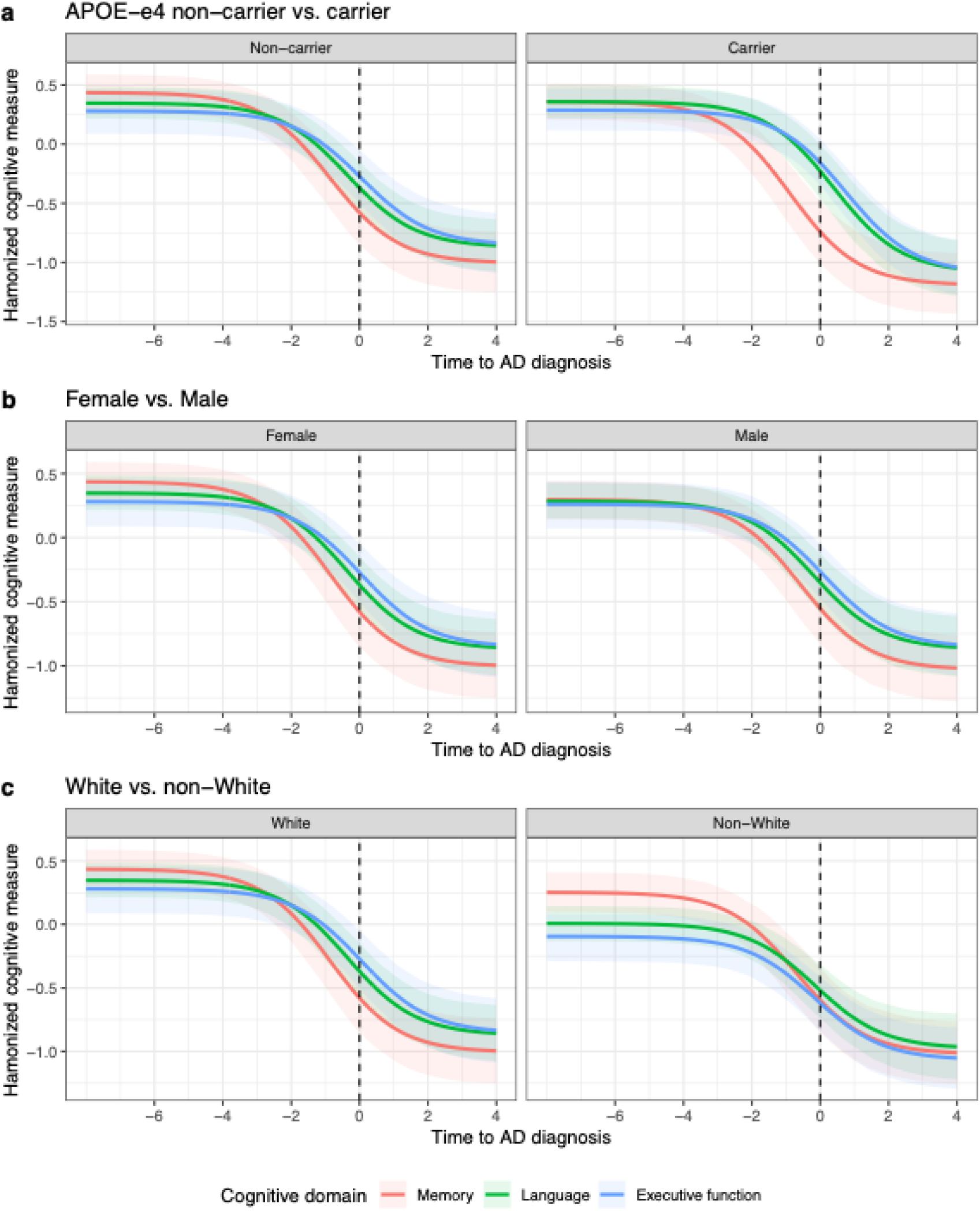
The estimated trajectories in the three cognitive domains (i.e., memory, language, and executive function) relative to the time to the diagnosis of AD in different subgroups. The shaded areas are 95% confidence intervals (CIs).

